## Supplementary File SF1: MNH_EPI _COS Acceptability Survey for "Acceptability and perceived barriers to adoption of the core outcome set for maternal and neonatal health research and surveillance during emerging and ongoing epidemic threats (MNH-EPI-COS). An online survey"

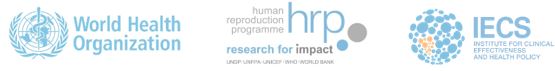

International online consultation on the acceptability of a core outcome set for maternal and neonatal health research and surveillance during emerging and ongoing epidemic threats

Between September 2023 and February 2024, the WHO conducted a four-stage modified Delphi study, involving two rounds of online surveys and two meetings, to reach a consensus on a core outcome set (COS) for maternal and neonatal health research and surveillance during epidemics.

Your contributions to the initial rounds of our Delphi process played a key role in shaping the consensus.

Following those rounds, and aligned with COS development guidelines, we continued the consensus process through in-depth discussions with a limited group of stakeholders. This group, representing the six WHO regions and diverse specialties, refined the outcomes, established definitions, and reached a final consensus.

We now invite you to participate in a new survey, designed to assess the acceptability of the COS and to gather feedback from the wider group of key stakeholders that participated in the online surveys.

Your insights will help us determine whether the online panels agree with and support the final COS obtained during the consensus process, and identify any areas that may require close monitoring during an implementation pilot. Additionally, we aim to assess the feasibility of collecting data for reporting these outcomes and to identify potential barriers to adopting this COS.

We deeply appreciate your continued involvement in this important work and look forward to your valuable feedback.

##### **Instructions**

The questionnaire is organized into four sections:

- Section 1: Participants’ characteristics
- Section 2: Acceptability of each outcome and feasibility of data collection
- Section 3: Overall acceptability of the COS
- Section 4: Barriers to implementation

Please complete the survey independently and anonymously to ensure that your responses reflect your personal views. Your participation is voluntary, and your responses will be kept confidential.

You can complete some sections and return later to finish, as your answers will be saved. We kindly ask that you complete the survey by October 7, 2024.

Thank you for your cooperation.

The survey should take approximately 30 - 40 minutes.

* 1. Consent to participate in the survey and Confidentiality Notice

By agreeing to participate in this survey, you will have access to the results of the modified Delphi study. This data is strictly confidential, and we require your commitment not to use it outside of this survey. Please select one option:

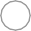
 Yes, I agree to participate and commit to maintaining the confidentiality of the results.

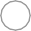
No, I do not give my consent to participate.

Results from previous rounds

The four-stage modified Delphi process supported the development of a final COS for maternal and neonatal health research and surveillance in light of emerging and ongoing epidemic threats (Box 1).

The COS is comprised of seven main maternal outcomes, eleven main neonatal outcomes, and eleven complementary outcomes, along with their definitions.

The main outcomes should be collected and reported in all studies during epidemics.

Complementary outcomes should be reported when relevant to the study type (e.g., epidemiological, product development, or post-authorization surveillance), the setting (considering resource availability and outcome validity), and the specific infectious disease outbreak. In research on emerging pathogens with unknown effects on maternal and neonatal health, researchers should strive to report all main and complementary outcomes or explain their reasoning for not reporting them.

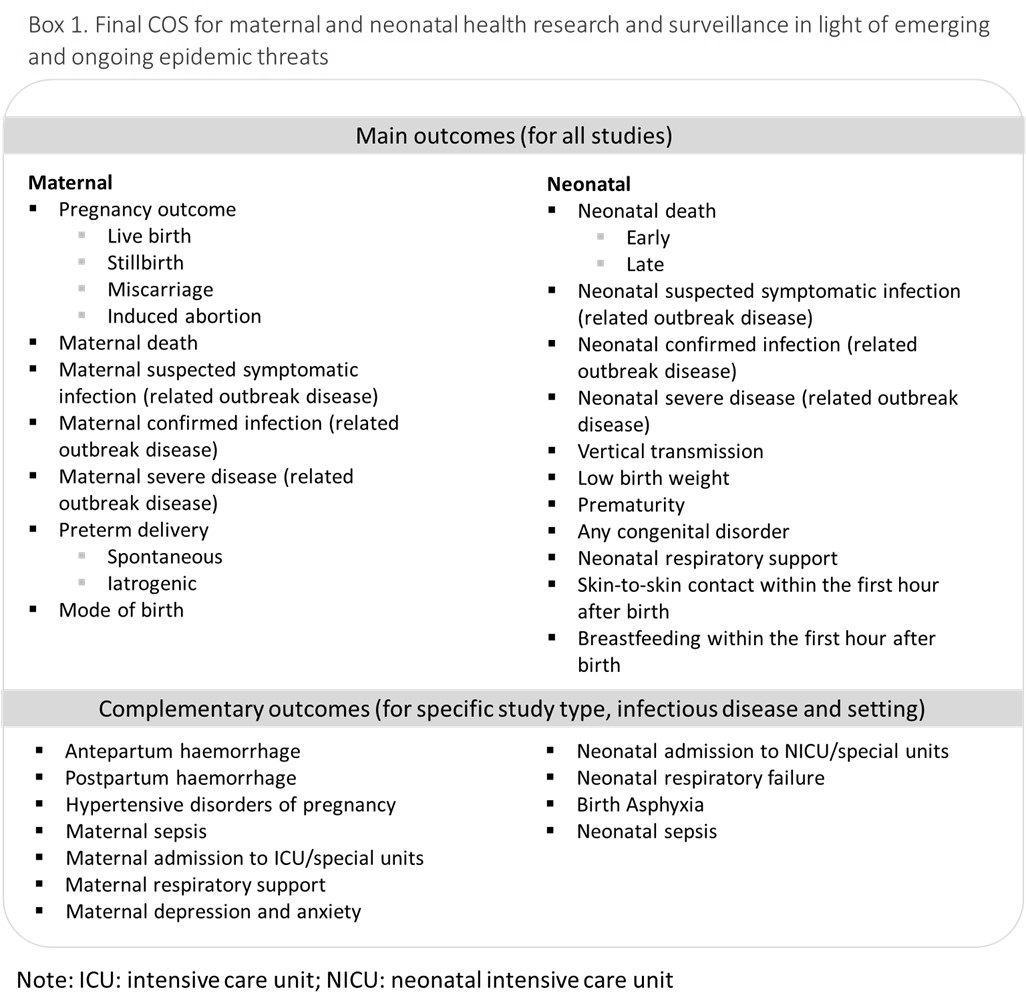

For detailed information on the study process, please read the [protocol paper](https://pubmed.ncbi.nlm.nih.gov/39175521/).

To review the results of the four-stage modified Delphi study, please refer to [this link](https://www.dropbox.com/scl/fi/mh3mpakt2028vs5yhyy2i/2.-Hyperlink_Summary-of-results_v3.0.pdf?rlkey=8mggfjtc175n2vsgdsvo2jp62&st=hi0082o5&dl=0).

Section 1: Participants characteristics

- 2. Which is your country of residence?

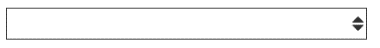

- 3. What is your gender?

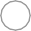
Male
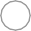
Female

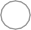
Non-binary
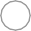
Other

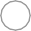
Prefer not to answer

- 4. What is your age?

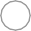
 <30

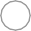
 30-39

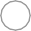
 40-49

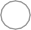
 50-59

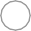
 >=60

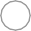
 Prefer not to answer

- 5. Which is the **main role** that describes you best?

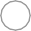
 Researcher

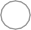
 Healthcare provider

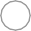
 Women/community/civil society representative

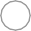
 Health service manager

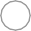
 Program manager
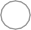
 Policy maker

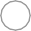
 Funder
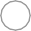
Regulator

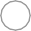
 Other (please specify)

- 6. Which **specialty or specialties** describe you best?

Maternal health

Epidemiology and public health

Neonatal and pediatric health

Patient advocacy

Infectious disease

Pharmacy/laboratory

Psychiatry/psychology/social work

Other (please specify)

**Some concepts to remember**

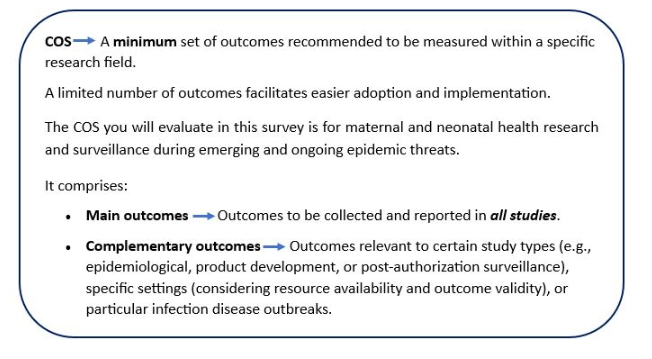

Section 2: Acceptability of each included outcome and feasibility of data collection

In the following section, we will ask for your opinion on:

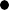
 The acceptability of each outcome included in the COS, their definitions and categorization, and the reasons for any disagreement.

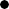
 The feasibility of collecting data to report these outcomes.

You will find details and questions about each outcome included in the COS. Each outcome includes its name, category, and definition as agreed upon in previous study rounds. Additionally, you may review notes and remarks for each outcome.

The proposed definitions align with the WHO and the International Classification of Diseases (ICD) standards, which are updated periodically.

First, all maternal outcomes, categorized as main or commentary, are presented. Later, neonatal outcomes will be presented.

### Main maternal outcomes

### Outcomes to be collected in all studies

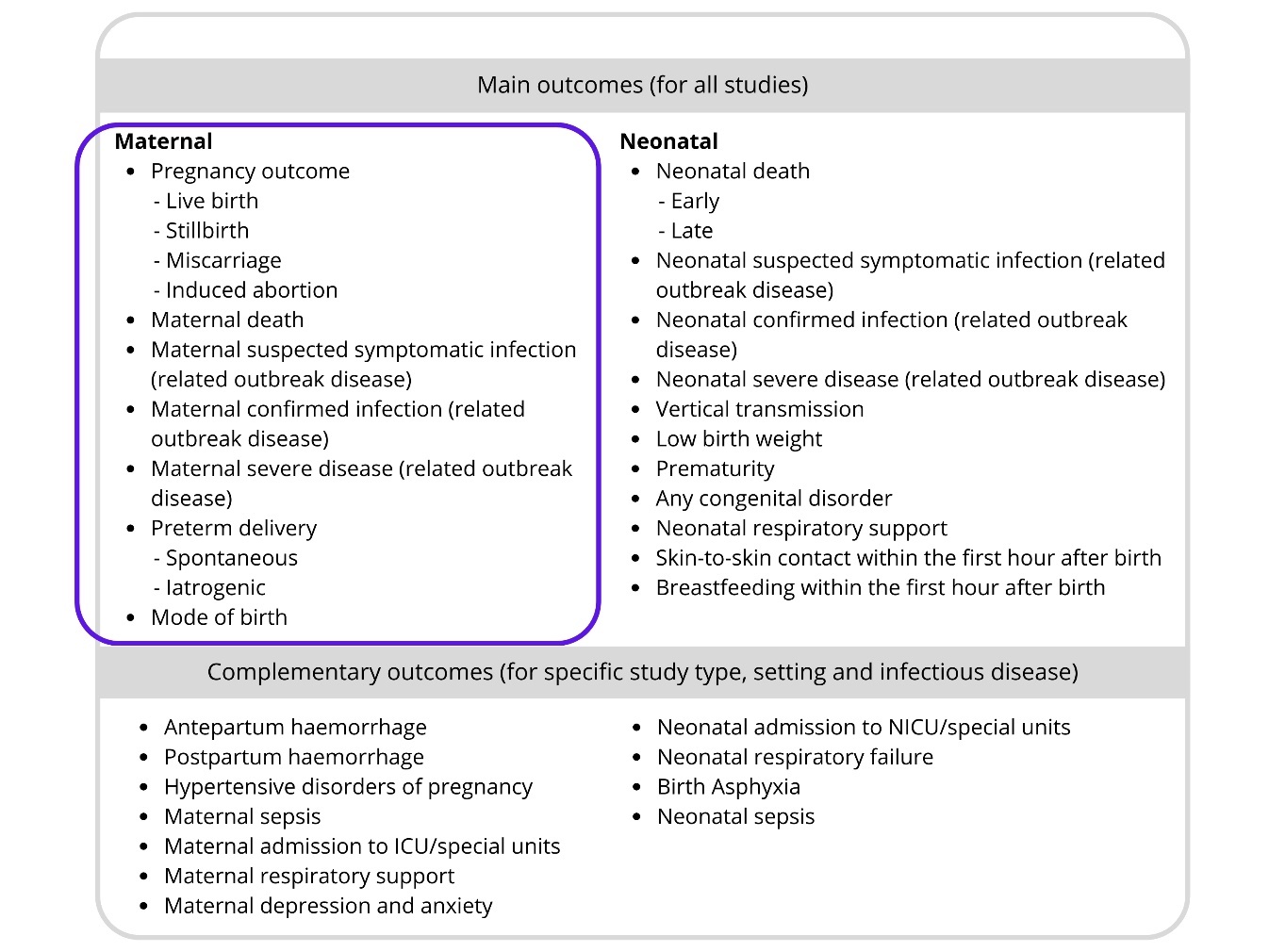

Pregnancy outcome

Category: Main outcome - to be collected in all studies

Definition

Results of conception and pregnancy, measured as:

- *Live birth* is an outcome of pregnancy, irrespective of the duration/gestation, where the newborn breathes or shows any other evidence of life–e.g, beating of the heart, pulsation of the umbilical cord or definite movement of voluntary muscles– whether the umbilical cord has been cut or the placenta is attached.
- *Stillbirth* is the complete expulsion or extraction from a woman of a fetus, following its death prior to the complete expulsion or extraction, at 22 or more completed weeks of gestation. When information on gestational age is unavailable use birthweight 500 grams or more as criteria.
- *Miscarriage* (also known as unintentional abortion or spontaneous abortion) is a spontaneous loss of pregnancy (i.e., embryo or fetus) before 22 completed
- weeks of gestation. When information on gestational age is not available, use birth weight of less than 500 grams as a criterion.
- *Induced abortion* is a complete expulsion or extraction from a woman of an embryo or a fetus (irrespective of the duration of the pregnancy) following a deliberate interruption of an ongoing pregnancy, which is not intended to result in a live birth.

You can access notes and remarks on this definition [here](https://www.dropbox.com/scl/fi/ax1jr5mx9jm5soasfi5r0/4.-Hyperlink-Maternal-outcomes-for-all-studies-Notes-Remarks.docx?rlkey=ivr4j1pq43uez24q98m9h5ci7&dl=0).

* 7. Pregnancy outcome

To what extent do you agree:

| Strongly  Disagree | Disagree | Neither Agree  Nor Disagree | Agree | Strongly Agree | *Unable to Assess* |
| --- | --- | --- | --- | --- | --- |

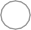

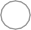

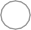

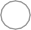

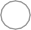

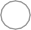

... to **include** this outcome as a main outcome?

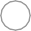

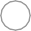

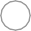

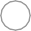

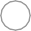

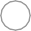
… with the proposed

**definitions**?

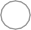

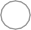

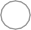

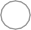

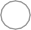

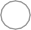

… that collecting this outcome in **observational and experimental** studies during **epidemics** is **feasible**?

If you DISAGREE or STRONGLY DISAGREE with one or more of the questions related to this outcome, please use this space below to provide your rationale

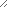

Maternal Death

Category: Main outcome - to be collected in all studies

Definition

Maternal death is defined as the death of a woman while pregnant or within 42 days of termination of pregnancy, irrespective of the duration and site of the pregnancy, from any cause related to or aggravated by the pregnancy or its management, but not from unintentional or incidental causes.

You can access notes and remarks on this definition [here](https://www.dropbox.com/scl/fi/ax1jr5mx9jm5soasfi5r0/4.-Hyperlink-Maternal-outcomes-for-all-studies-Notes-Remarks.docx?rlkey=ivr4j1pq43uez24q98m9h5ci7&dl=0).

* 8. Maternal death

To what extent do you agree:

| Strongly  Disagree | Disagree | Neither Agree  Nor Disagree | Agree | Strongly Agree | *Unable to Assess* |
| --- | --- | --- | --- | --- | --- |

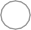

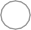

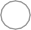

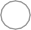

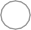

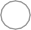

... to **include** this outcome as a main outcome?

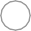

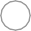

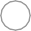

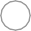

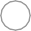

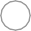
… with the proposed

**definition**?

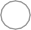

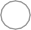

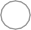

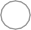

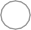

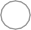

… that collecting this outcome in **observational and experimental** studies during **epidemics** is **feasible**?

If you DISAGREE or STRONGLY DISAGREE with one or more of the questions related to this outcome, please use this space below to provide your rationale

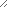

Maternal suspected symptomatic infection

Category: Main outcome - to be collected in all studies

Definition

A woman who meets clinical criteria (signs and symptoms) for the disease under investigation while pregnant or within 42 days after birth.

You can access notes and remarks on this definition [here](https://www.dropbox.com/scl/fi/ax1jr5mx9jm5soasfi5r0/4.-Hyperlink-Maternal-outcomes-for-all-studies-Notes-Remarks.docx?rlkey=ivr4j1pq43uez24q98m9h5ci7&dl=0).

* 9. Maternal suspected symptomatic infection

To what extent do you agree:

| Strongly  Disagree | Disagree | Neither Agree  Nor Disagree | Agree | Strongly Agree | *Unable to Assess* |
| --- | --- | --- | --- | --- | --- |

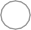

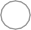

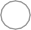

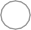

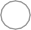

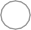

... to **include** this outcome as a main outcome?

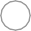

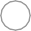

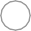

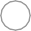

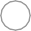

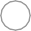
… with the proposed

**definition**?

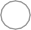

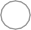

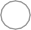

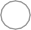

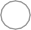

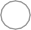

… that collecting this outcome in **observational and experimental** studies during **epidemics** is **feasible**?

If you DISAGREE or STRONGLY DISAGREE with one or more of the questions related to this outcome, please use this space below to provide your rationale

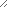

Maternal confirmed infection (related outbreak disease)

Category: Main outcome - to be collected in all studies

Definition

A woman with a positive and confirmatory laboratory or pathology result while pregnant or within 42 days of end of pregnancy, regardless of clinical OR epidemiological criteria for the disease under investigation, and vital status.

You can access notes and remarks on this definition [here](https://www.dropbox.com/scl/fi/ax1jr5mx9jm5soasfi5r0/4.-Hyperlink-Maternal-outcomes-for-all-studies-Notes-Remarks.docx?rlkey=ivr4j1pq43uez24q98m9h5ci7&dl=0)

* 10. Maternal confirmed infection (related outbreak disease)

To what extent do you agree:

| Strongly  Disagree | Disagree | Neither Agree  Nor Disagree | Agree | Strongly Agree | *Unable to Assess* |
| --- | --- | --- | --- | --- | --- |

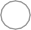

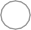

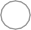

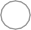

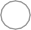

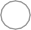

... to **include** this outcome as a main outcome?

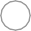

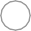

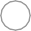

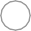

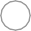

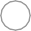
… with the proposed

**definition**?

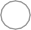

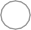

… that collecting this outcome in **observational and experimental** studies during **epidemics** is **feasible**?

If you DISAGREE or STRONGLY DISAGREE with one or more of the questions related to this outcome, please use this space below to provide your rationale

Maternal severe disease (related outbreak disease)

Category: Main outcome - to be collected in all studies

Definition

A woman presenting while pregnant or within 42 days of end of pregnancy with illness (specific to the disease under investigation) that results in acute physiological instability (abnormal physiological parameters or vital organ dysfunction or failure) or a clinical support requirement (such as hospitalization, intensive care or high-dependency unit or time-sensitive intervention) to prevent clinical deterioration, disability or death.

You can access notes and remarks on this definition [here](https://www.dropbox.com/scl/fi/ax1jr5mx9jm5soasfi5r0/4.-Hyperlink-Maternal-outcomes-for-all-studies-Notes-Remarks.docx?rlkey=ivr4j1pq43uez24q98m9h5ci7&dl=0).

* 11. Maternal severe disease (related outbreak disease)

To what extent do you agree:

| Strongly  Disagree | Disagree | Neither Agree  Nor Disagree | Agree | Strongly Agree | *Unable to Assess* |
| --- | --- | --- | --- | --- | --- |

... to **include** this outcome as a main outcome?

… with the proposed

**definitions**?

… that collecting this outcome in **observational and experimental** studies during **epidemics** is **feasible**?

If you DISAGREE or STRONGLY DISAGREE with one or more of the questions related to this outcome, please use this space below to provide your rationale

Spontaneous/iatrogenic preterm birth

Category: Main outcome - to be collected in all studies

Definition

Preterm is defined as babies born alive before 37 weeks of pregnancy are completed.

- *Spontaneous preterm birth* is defined as the spontaneous onset of labour and delivery of a live born between 22 and 36 completed weeks.
- *Iatrogenic preterm birth* (also known as medically indicated for fetal and/or maternal interest), is defined as a live born between 22 and 36 completed weeks after induction of labour or a caesarean before labour.

You can access notes and remarks on this definition [here](https://www.dropbox.com/scl/fi/ax1jr5mx9jm5soasfi5r0/4.-Hyperlink-Maternal-outcomes-for-all-studies-Notes-Remarks.docx?dl=0&rlkey=4kax4ikwzqmbi76kp23rx513v).

* 12. Spontaneous/iatrogenic preterm birth

To what extent do you agree:

| Strongly  Disagree | Disagree | Neither Agree  Nor Disagree | Agree | Strongly Agree | *Unable to Assess* |
| --- | --- | --- | --- | --- | --- |

... to **include** this outcome as a main outcome?

… with the proposed

**definitions**?

… that collecting this outcome in **observational and experimental** studies during **epidemics** is **feasible**?

If you DISAGREE or STRONGLY DISAGREE with one or more of the questions related to this outcome, please use this space below to provide your rationale

Mode of birth

Category: Main outcome - to be collected in all studies

Definition

Parturition of a newborn from the uterus via spontaneous vaginal birth, assisted vaginal birth (vacuum or forceps), elective caesarean section, or emergency caesarean section.

You can access notes and remarks on this definition [here](https://www.dropbox.com/scl/fi/ax1jr5mx9jm5soasfi5r0/4.-Hyperlink-Maternal-outcomes-for-all-studies-Notes-Remarks.docx?rlkey=ivr4j1pq43uez24q98m9h5ci7&dl=0).

* 13. Mode of birth

To what extent do you agree:

| Strongly  Disagree | Disagree | Neither Agree  Nor Disagree | Agree | Strongly Agree | *Unable to Assess* |
| --- | --- | --- | --- | --- | --- |

... to **include** this outcome as a main outcome?

… with the proposed

**definition**?

… that collecting this outcome in **observational and experimental** studies during **epidemics** is **feasible**?

If you DISAGREE or STRONGLY DISAGREE with one or more of the questions related to this outcome, please use this space below to provide your rationale

### Maternal & pregnancy complementary outcomes

### Outcomes relevant to specific study types, settings, or particular disease outbreaks

Antepartum hemorrhage

Category: Complementary outcome - relevant to specific study types, settings, or particular disease outbreaks

Definition

Vaginal bleeding after 22 weeks of pregnancy or during labour before giving birth.

You can access the discussion remarks and notes on the definitions [here](https://www.dropbox.com/scl/fi/iy0varqjoqmtpkg6q5717/4.-Hyperlink-Maternal-complementary-outcomes-Notes-Remarks.docx?dl=0&rlkey=30zwyrjf66lmx8bif3he2c15b).

* 14. Antepartum hemorrhage

To what extent do you agree:

| Strongly  Disagree | Disagree | Neither Agree  Nor Disagree | Agree | Strongly Agree | *Unable to Assess* |
| --- | --- | --- | --- | --- | --- |

... to include this outcome as a **complementary outcome** - to be collected if relevant to specific study types, settings, or particular disease outbreaks?

… with the proposed

**definition**?

… that **collecting** this outcome in **observational and experimental** studies during **epidemics** is feasible?

If you DISAGREE or STRONGLY DISAGREE with one or more of the questions related to this outcome, please use this space below to provide your rationale

Postpartum hemorrhage

Category: Complementary outcome - relevant to specific study types, settings, or particular disease outbreaks

Definition

A blood loss of 500 ml or more within 24 hours after birth.

You can access the discussion remarks and notes on the definitions [here](https://www.dropbox.com/scl/fi/iy0varqjoqmtpkg6q5717/4.-Hyperlink-Maternal-complementary-outcomes-Notes-Remarks.docx?rlkey=30zwyrjf66lmx8bif3he2c15b&dl=0).

* 15. Postpartum hemorrhage

To what extent do you agree:

| Strongly  Disagree | Disagree | Neither Agree  Nor Disagree | Agree | Strongly Agree | *Unable to Assess* |
| --- | --- | --- | --- | --- | --- |

... to include this outcome as a **complementary outcome** - to be collected if relevant to specific study types, settings, or particular disease outbreaks?

… with the proposed

**definition**?

… that **collecting** this outcome in **observational and experimental** studies during **epidemics** is feasible?

If you DISAGREE or STRONGLY DISAGREE with one or more of the questions related to this outcome, please use this space below to provide your rationale

**Hypertensive disorders of pregnancy**

**Category: Complementary outcome - relevant to specific study types, settings, or particular disease outbreaks**

Definition

A hypertensive disorder newly diagnosed after 20 weeks´ gestation or before 1 week postpartum, characterized by systolic blood pressure greater than 140mmHg and/or a diastolic blood pressure greater or equal to 90mmHg on two occasions, 4 hours or more apart.

You can access the discussion remarks and notes on the definitions [here](https://www.dropbox.com/scl/fi/iy0varqjoqmtpkg6q5717/4.-Hyperlink-Maternal-complementary-outcomes-Notes-Remarks.docx?rlkey=30zwyrjf66lmx8bif3he2c15b&dl=0).

* 16. Hypertensive disorders of pregnancy

To what extent do you agree:

| Strongly  Disagree | Disagree | Neither Agree  Nor Disagree | Agree | Strongly Agree | *Unable to Assess* |
| --- | --- | --- | --- | --- | --- |

... to include this outcome as a **complementary outcome** - to be collected if relevant to specific study types, settings, or particular disease outbreaks?

… with the proposed

**definition**?

… that **collecting** this outcome in **observational and experimental** studies during **epidemics** is feasible?

If you DISAGREE or STRONGLY DISAGREE with one or more of the questions related to this outcome, please use this space below to provide your rationale

Maternal sepsis

Category: Complementary outcome - relevant to specific study types, settings, or particular disease outbreaks

Definition

A life-threatening condition defined as organ dysfunction resulting from infection during pregnancy, childbirth, post-abortion, or post-partum period.

You can access the discussion remarks and notes on the definitions [here](https://www.dropbox.com/scl/fi/iy0varqjoqmtpkg6q5717/4.-Hyperlink-Maternal-complementary-outcomes-Notes-Remarks.docx?rlkey=30zwyrjf66lmx8bif3he2c15b&dl=0).

* 17. Maternal sepsis

To what extent do you agree:

| Strongly  Disagree | Disagree | Neither Agree  Nor Disagree | Agree | Strongly Agree | *Unable to Assess* |
| --- | --- | --- | --- | --- | --- |

... to include this outcome as a **complementary outcome** - to be collected if relevant to specific study types, settings, or particular disease outbreaks?

… with the proposed

**definition**?

… that **collecting** this outcome in **observational and experimental** studies during **epidemics** is feasible?

If you DISAGREE or STRONGLY DISAGREE with one or more of the questions related to this outcome, please use this space below to provide your rationale

**Maternal admission to intensive care unit / special unit**

**Category: Complementary outcome - relevant to specific study types, settings, or particular disease outbreaks**

Definition

Admission to an intensive care unit or a unit that provides 24-hour monitoring and vital support at any point during pregnancy through 42 days after pregnancy (postpartum, post-abortion/miscarriage) for any obstetric indication, outbreak disease or other non-obstetric indications.

You can access the discussion remarks and notes on the definitions [here.](https://www.dropbox.com/scl/fi/iy0varqjoqmtpkg6q5717/4.-Hyperlink-Maternal-complementary-outcomes-Notes-Remarks.docx?rlkey=30zwyrjf66lmx8bif3he2c15b&dl=0)

* 18. Maternal admission to intensive care unit / special unit

To what extent do you agree:

| Strongly  Disagree | Disagree | Neither Agree  Nor Disagree | Agree | Strongly Agree | *Unable to Assess* |
| --- | --- | --- | --- | --- | --- |

... to include this outcome as a **complementary outcome** - to be collected if relevant to specific study types, settings, or particular disease outbreaks?

… with the proposed

**definition**?

… that **collecting** this outcome in **observational and experimental** studies during **epidemics** is feasible?

If you DISAGREE or STRONGLY DISAGREE with one or more of the questions related to this outcome, please use this space below to provide your rationale

**Maternal respiratory support**

**Category: Complementary outcome - relevant to specific study types, settings, or particular disease outbreaks**

Definition

Any respiratory support technique with any level of pressure via any interface, during pregnancy, -labour and 42 days after pregnancy, not related to anaesthesia during delivery (postpartum/postabortion).

You can access the discussion remarks and notes on the definitions [here](https://www.dropbox.com/scl/fi/iy0varqjoqmtpkg6q5717/4.-Hyperlink-Maternal-complementary-outcomes-Notes-Remarks.docx?dl=0&rlkey=30zwyrjf66lmx8bif3he2c15b).

* 19. Maternal respiratory support

To what extent do you agree:

| Strongly  Disagree | Disagree | Neither Agree  Nor Disagree | Agree | Strongly Agree | *Unable to Assess* |
| --- | --- | --- | --- | --- | --- |

... to include this outcome as a **complementary outcome** - to be collected if relevant to specific study types, settings, or particular disease outbreaks?

… with the proposed

**definition**?

… that **collecting** this outcome in **observational and experimental** studies during **epidemics** is feasible?

If you DISAGREE or STRONGLY DISAGREE with one or more of the questions related to this outcome, please use this space below to provide your rationale

**Maternal symptoms of depression and anxiety**

**Category: Complementary outcome - relevant to specific study types, settings, or particular disease outbreaks**

Definition

*Depressive disorders* are characterized by sadness, loss of interest or pleasure, feelings of guilt or low self-worth, disturbed sleep or appetite, feelings of tiredness, and poor concentration.

*Anxiety disorders* refer to a group of mental disorders characterized by feelings of anxiety and fear, including generalized anxiety disorder (GAD), panic disorder, phobias, social anxiety disorder, obsessive-compulsive disorder (OCD) and post- traumatic stress disorder (PTSD).

You can access the discussion remarks and notes on the definitions [here](https://www.dropbox.com/scl/fi/iy0varqjoqmtpkg6q5717/4.-Hyperlink-Maternal-complementary-outcomes-Notes-Remarks.docx?dl=0&rlkey=30zwyrjf66lmx8bif3he2c15b).

* 20. Maternal symptoms of depression and anxiety

To what extent do you agree:

| Strongly  Disagree | Disagree | Neither Agree  Nor Disagree | Agree | Strongly Agree | *Unable to Assess* |
| --- | --- | --- | --- | --- | --- |

... to include this outcome as a **complementary outcome** - to be collected if relevant to specific study types, settings, or particular disease outbreaks?

… with the proposed

**definitions**?

… that **collecting** this outcome in **observational and experimental** studies during **epidemics** is feasible?

If you DISAGREE or STRONGLY DISAGREE with one or more of the questions related to this outcome, please use this space below to provide your rationale

### Main neonatal outcomes

### Outcomes to be collected in all studies

#### Neonatal death

Category: Main outcome - to be collected in all studies

Definition

A death following a live birth, with 22 or more completed weeks of gestation (or 500 g or more) occurring from time of birth to 28 days after birth (note day 1 is the day of birth).

You can access the discussion remarks and notes on the definitions [here](https://www.dropbox.com/scl/fi/xrafsy2ygv47fq4k3bgir/5.-Hyperlink-Neonatal-outcomes-for-all-studies-Notes-Remarks.docx?rlkey=fcjryqyez5r105awfifzy8jfk&dl=0).

* 21. Neonatal death

To what extent do you agree:

| Strongly  Disagree | Disagree | Neither Agree  Nor Disagree | Agree | Strongly Agree | *Unable to Assess* |
| --- | --- | --- | --- | --- | --- |

... to **include** this outcome as a main outcome?

… with the proposed

**definition**?

… that collecting this outcome in **observational and experimental** studies during **epidemics** is **feasible**?

If you DISAGREE or STRONGLY DISAGREE with one or more of the questions related to this outcome, please use this space below to provide your rationale

#### Neonatal suspected symptomatic infection (related outbreak disease)

Category: Main outcome - to be collected in all studies

Definition

A newborn (1-28 days after birth) who meets clinical criteria (signs and symptoms) associated with a disease under investigation, including antenatal and postnatal infection.

You can access the discussion remarks and notes on the definitions [here](https://www.dropbox.com/scl/fi/xrafsy2ygv47fq4k3bgir/5.-Hyperlink-Neonatal-outcomes-for-all-studies-Notes-Remarks.docx?dl=0&rlkey=fcjryqyez5r105awfifzy8jfk).

* 22. Neonatal suspected symptomatic infection (related outbreak disease)

To what extent do you agree:

| Strongly  Disagree | Disagree | Neither Agree  Nor Disagree | Agree | Strongly Agree | *Unable to Assess* |
| --- | --- | --- | --- | --- | --- |

... to **include** this outcome as a main outcome?

… with the proposed

**definition**?

… that collecting this outcome in **observational and experimental** studies during **epidemics** is **feasible**?

If you DISAGREE or STRONGLY DISAGREE with one or more of the questions related to this outcome, please use this space below to provide your rationale

#### Neonatal confirmed infection (related outbreak disease)

Category: Main outcome - to be collected in all studies

Definition

A newborn (1-28 days after birth) with a positive and confirmatory laboratory or pathology result (recognized pathogen identified using a validated method), regardless of clinical criteria OR epidemiological criteria (related outbreak in investigation), antenatal or postnatal acquisition, and vital status.

You can access the discussion remarks and notes on the definitions [here](https://www.dropbox.com/scl/fi/xrafsy2ygv47fq4k3bgir/5.-Hyperlink-Neonatal-outcomes-for-all-studies-Notes-Remarks.docx?dl=0&rlkey=fcjryqyez5r105awfifzy8jfk).

* 23. Neonatal confirmed infection (related outbreak disease)

To what extent do you agree:

| Strongly  Disagree | Disagree | Neither Agree  Nor Disagree | Agree | Strongly Agree | *Unable to Assess* |
| --- | --- | --- | --- | --- | --- |

... to **include** this outcome as a main outcome?

… with the proposed

**definition**?

… that collecting this outcome in **observational and experimental** studies during **epidemics** is **feasible**?

If you DISAGREE or STRONGLY DISAGREE with one or more of the questions related to this outcome, please use this space below to provide your rationale

#### Severe/critical disease (related outbreak disease)

Category: Main outcome - to be collected in all studies

Definition

A newborn (1- 28 days) with an antenatally or postnatally acquired illness (specific to the outbreak under investigation) that results in acute physiological instability (abnormal physiological parameters or vital organ dysfunction or failure) or a clinical support requirement (such as hospitalization, admission to NICU, intensive care or high-dependency unit or time-sensitive intervention) to prevent further clinical deterioration, disability, or death.

You can access the discussion remarks and notes on the definitions [here](https://www.dropbox.com/scl/fi/xrafsy2ygv47fq4k3bgir/5.-Hyperlink-Neonatal-outcomes-for-all-studies-Notes-Remarks.docx?rlkey=fcjryqyez5r105awfifzy8jfk&dl=0).

* 24. Severe/critical disease (related outbreak disease)

To what extent do you agree:

| Strongly  Disagree | Disagree | Neither Agree  Nor Disagree | Agree | Strongly Agree | *Unable to Assess* |
| --- | --- | --- | --- | --- | --- |

... to **include** this outcome as a main outcome?

… with the proposed

**definition**?

… that collecting this outcome in **observational and experimental** studies during **epidemics** is **feasible**?

If you DISAGREE or STRONGLY DISAGREE with one or more of the questions related to this outcome, please use this space below to provide your rationale

#### Vertical transmission

Category: Main outcome - to be collected in all studies

Definition

Transmission of pathogen from a parent to the fetus or baby during pregnancy (in utero), intrapartum by exposure to blood and secretions, and by exposure after birth via breast milk.

You can access the discussion remarks and notes on the definitions [here](https://www.dropbox.com/scl/fi/xrafsy2ygv47fq4k3bgir/5.-Hyperlink-Neonatal-outcomes-for-all-studies-Notes-Remarks.docx?rlkey=fcjryqyez5r105awfifzy8jfk&dl=0).

* 25. Vertical transmission

To what extent do you agree:

| Strongly  Disagree | Disagree | Neither Agree  Nor Disagree | Agree | Strongly Agree | *Unable to Assess* |
| --- | --- | --- | --- | --- | --- |

... to **include** this outcome as a main outcome?

… with the proposed

**definition**?

… that collecting this outcome in **observational and experimental** studies during **epidemics** is **feasible**?

If you DISAGREE or STRONGLY DISAGREE with one or more of the questions related to this outcome, please use this space below to provide your rationale

#### Low birthweight

Category: Main outcome - to be collected in all studies

Definition

A live born with weight less than 2500 g at birth. Sub-categories include:

- Extremely low birth weight (<1000 g)
- Very low birth weight (1000 - 1499 g)
- Low birth weight (1500 - 2499 g)

You can access the discussion remarks and notes on the definitions [here](https://www.dropbox.com/scl/fi/xrafsy2ygv47fq4k3bgir/5.-Hyperlink-Neonatal-outcomes-for-all-studies-Notes-Remarks.docx?dl=0&rlkey=fcjryqyez5r105awfifzy8jfk).

* 26. Low birthweight

To what extent do you agree:

| Strongly  Disagree | Disagree | Neither Agree  Nor Disagree | Agree | Strongly Agree | *Unable to Assess* |
| --- | --- | --- | --- | --- | --- |

... to **include** this outcome as a main outcome?

… with the proposed

**definition**?

… that collecting this outcome in **observational and experimental** studies during **epidemics** is **feasible**?

If you DISAGREE or STRONGLY DISAGREE with one or more of the questions related to this outcome, please use this space below to provide your rationale

#### Prematurity

Category: Main outcome - to be collected in all studies

Definition

Babies born alive greater than or equal to 22+0 and less than 37 completed weeks of gestation. Sub-categories include:

- Extremely preterm (22+0 w to 27+6)
- Very preterm (28+0 w to 31+6)
- Moderately preterm (32+0 w to 33+6)
- Late preterm (from 34+0 weeks to 36+6)

You can access the discussion remarks and notes on the definitions [here](https://www.dropbox.com/scl/fi/xrafsy2ygv47fq4k3bgir/5.-Hyperlink-Neonatal-outcomes-for-all-studies-Notes-Remarks.docx?dl=0&rlkey=fcjryqyez5r105awfifzy8jfk).

* 27. Prematurity

To what extent do you agree:

| Strongly  Disagree | Disagree | Neither Agree  Nor Disagree | Agree | Strongly Agree | *Unable to Assess* |
| --- | --- | --- | --- | --- | --- |

... to **include** this outcome as a main outcome?

… with the proposed

**definition**?

… that collecting this outcome in **observational and experimental** studies during **epidemics** is **feasible**?

If you DISAGREE or STRONGLY DISAGREE with one or more of the questions related to this outcome, please use this space below to provide your rationale

#### Any congenital anomaly

Category: Main outcome - to be collected in all studies

Definition

Any structural or functional anomalies that develop in utero, and may be identified before, at birth, or after birth.

You can access the discussion remarks and notes on the definitions [here.](https://www.dropbox.com/scl/fi/xrafsy2ygv47fq4k3bgir/5.-Hyperlink-Neonatal-outcomes-for-all-studies-Notes-Remarks.docx?dl=0&rlkey=fcjryqyez5r105awfifzy8jfk)

* 28. Any congenital anomaly

To what extent do you agree:

| Strongly  Disagree | Disagree | Neither Agree  Nor Disagree | Agree | Strongly Agree | *Unable to Assess* |
| --- | --- | --- | --- | --- | --- |

... to **include** this outcome as a main outcome?

… with the proposed

**definition**?

… that collecting this outcome in **observational and experimental** studies during **epidemics** is **feasible**?

If you DISAGREE or STRONGLY DISAGREE with one or more of the questions related to this outcome, please use this space below to provide your rationale

#### Neonatal respiratory support

Category: Main outcome - to be collected in all studies

Definition

Any respiratory support technique to a neonate between day 1 and 28 after birth with any level of pressure via any interface, not related to anaesthesia.

You can access the discussion remarks and notes on the definitions [here](https://www.dropbox.com/scl/fi/xrafsy2ygv47fq4k3bgir/5.-Hyperlink-Neonatal-outcomes-for-all-studies-Notes-Remarks.docx?dl=0&rlkey=fcjryqyez5r105awfifzy8jfk).

* 29. Neonatal respiratory support

To what extent do you agree:

| Strongly  Disagree | Disagree | Neither Agree  Nor Disagree | Agree | Strongly Agree | *Unable to Assess* |
| --- | --- | --- | --- | --- | --- |

... to **include** this outcome as a main outcome?

… with the proposed

**definition**?

… that collecting this outcome in **observational and experimental** studies during **epidemics** is **feasible**?

If you DISAGREE or STRONGLY DISAGREE with one or more of the questions related to this outcome, please use this space below to provide your rationale

#### Skin-to-skin contact during the first hour after birth

Category: Main outcome - to be collected in all studies

Definition

A newborn without complications is kept in skin-to-skin contact with her/his mother, placed prone on the mother’s abdomen or chest in direct ventral-to-ventral skin-to- skin contact for at least an hour or until after the first feed.

You can access the discussion remarks and notes on the definitions [here](https://www.dropbox.com/scl/fi/xrafsy2ygv47fq4k3bgir/5.-Hyperlink-Neonatal-outcomes-for-all-studies-Notes-Remarks.docx?dl=0&rlkey=fcjryqyez5r105awfifzy8jfk).

* 30. Skin-to-skin contact during the first hour after birth

To what extent do you agree:

| Strongly  Disagree | Disagree | Neither Agree  Nor Disagree | Agree | Strongly Agree | *Unable to Assess* |
| --- | --- | --- | --- | --- | --- |

... to **include** this outcome as a main outcome?

… with the proposed

**definition**?

… that collecting this outcome in **observational and experimental** studies during **epidemics** is **feasible**?

If you DISAGREE or STRONGLY DISAGREE with one or more of the questions related to this outcome, please use this space below to provide your rationale

#### Breastfeeding within one hour of birth

Category: Main outcome - to be collected in all studies

Definition

Newborn who is put to the breast during as soon as possible after birth and within one hour of birth, when they are clinically stable, and the mother and baby are ready.

You can access the discussion remarks and notes on the definitions [here](https://www.dropbox.com/scl/fi/xrafsy2ygv47fq4k3bgir/5.-Hyperlink-Neonatal-outcomes-for-all-studies-Notes-Remarks.docx?dl=0&rlkey=fcjryqyez5r105awfifzy8jfk).

* 31. Breastfeeding within one hour of birth

To what extent do you agree:

| Strongly  Disagree | Disagree | Neither Agree  Nor Disagree | Agree | Strongly Agree | *Unable to Assess* |
| --- | --- | --- | --- | --- | --- |

... to **include** this outcome as a main outcome?

… with the proposed

**definition**?

… that collecting this outcome in **observational and experimental** studies during **epidemics** is **feasible**?

If you DISAGREE or STRONGLY DISAGREE with one or more of the questions related to this outcome, please use this space below to provide your rationale

### Neonatal complementary outcomes

### Outcomes relevant to specific study types, settings, or

**particular disease outbreaks**

**Neonatal admission to the intensive care unit/other special units**

**Category: Complementary outcome - relevant to specific study types, settings, or particular disease outbreaks**

Definition

Admission of a neonate in the first 28 days after birth to an intensive care unit or a unit that provides 24-hour monitoring and vital support for any indications.

You can access the discussion remarks and notes on the definitions [here](https://www.dropbox.com/scl/fi/nmokygrj21p6466547rct/5.-Hyperlink-Neonatal-complementary-outcomes-Notes-Remarks.pdf?rlkey=9ezhtv9up0nccsmngubrbtn20&dl=0).

* 32. Neonatal admission to the intensive care unit/other special units

To what extent do you agree:

| Strongly  Disagree | Disagree | Neither Agree  Nor Disagree | Agree | Strongly Agree | *Unable to Assess* |
| --- | --- | --- | --- | --- | --- |

... to include this outcome as a **complementary outcome** - to be collected if relevant to specific study types, settings, or particular disease outbreaks?

… with the proposed

**definition**?

… that **collecting** this outcome in **observational and experimental** studies during **epidemics** is feasible?

If you DISAGREE or STRONGLY DISAGREE with one or more of the questions related to this outcome, please use this space below to provide your rationale

**Neonatal respiratory failure**

**Category: Complementary outcome - relevant to specific study types, settings, or particular disease outbreaks**

Definition

Acute onset of respiratory failure in a newborn (any gestational age or birthweight), in the first 28 days after birth, from any cause. It is recognized as one or more signs of increased work of breathing (such as tachypnoea, nasal flaring, chest retractions, or grunting) with or without cyanosis.

You can access the discussion remarks and notes on the definitions [here](https://www.dropbox.com/scl/fi/nmokygrj21p6466547rct/5.-Hyperlink-Neonatal-complementary-outcomes-Notes-Remarks.pdf?rlkey=9ezhtv9up0nccsmngubrbtn20&dl=0).

* 33. Neonatal respiratory failure

To what extent do you agree:

| Strongly  Disagree | Disagree | Neither Agree  Nor Disagree | Agree | Strongly Agree | *Unable to Assess* |
| --- | --- | --- | --- | --- | --- |

... to include this outcome as a **complementary outcome** - to be collected if relevant to specific study types, settings, or particular disease outbreaks?

… with the proposed

**definition**?

… that **collecting** this outcome in **observational and experimental** studies during **epidemics** is feasible?

If you DISAGREE or STRONGLY DISAGREE with one or more of the questions related to this outcome, please use this space below to provide your rationale

Birth asphyxia

Category: Complementary outcome - relevant to specific study types, settings, or particular disease outbreaks

Definition

Birth asphyxia is the inability of a newborn to start and sustain breathing immediately after birth, characterized by a low Apgar score and/or metabolic acidosis.

It is classified in:

- Mild and moderate: Normal respiration not established within one minute, but heart rate 100 per minute or above, some muscle tone present, some response to stimulation. Apgar score 4-7 at 5 minutes and/or pH <7.2 on the umbilical cord arterial blood sample
- Severe: Pulse less than 100 per minute at birth and falling or steady, respiration absent or gasping, colour poor, tone absent. Apgar score 0-3 at 5 minutes. And/or also characterized by profound metabolic acidosis, pH < 7.0 on the umbilical cord arterial blood sample.

You can access the discussion remarks and notes on the definitions [here](https://www.dropbox.com/scl/fi/nmokygrj21p6466547rct/5.-Hyperlink-Neonatal-complementary-outcomes-Notes-Remarks.pdf?rlkey=9ezhtv9up0nccsmngubrbtn20&dl=0).

* 34. Birth asphyxia

To what extent do you agree:

| Strongly  Disagree | Disagree | Neither Agree  Nor Disagree | Agree | Strongly Agree | *Unable to Assess* |
| --- | --- | --- | --- | --- | --- |

... to include this outcome as a **complementary outcome** - to be collected if relevant to specific study types, settings, or particular disease outbreaks?

… with the proposed

**definition**?

… that **collecting** this outcome in **observational and experimental** studies during **epidemics** is feasible?

If you DISAGREE or STRONGLY DISAGREE with one or more of the questions related to this outcome, please use this space below to provide your rationale

Neonatal sepsis

Category: Complementary outcome - relevant to specific study types, settings, or particular disease outbreaks

Definition

A condition affecting fetuses or newborns, that is (or suspected to be) caused by a maternal infection (acquired in utero or during birth) with a bacterial, viral, fungal, or parasitic source.

You can access the discussion remarks and notes on the definitions [here](https://www.dropbox.com/scl/fi/nmokygrj21p6466547rct/5.-Hyperlink-Neonatal-complementary-outcomes-Notes-Remarks.pdf?rlkey=9ezhtv9up0nccsmngubrbtn20&dl=0).

* 35. Neonatal sepsis

To what extent do you agree:

| Strongly  Disagree | Disagree | Neither Agree  Nor Disagree | Agree | Strongly Agree | *Unable to Assess* |
| --- | --- | --- | --- | --- | --- |

... to include this outcome as a **complementary outcome** - to be collected if relevant to specific study types, settings, or particular disease outbreaks?

… with the proposed

**definition**?

… that **collecting** this outcome in **observational and experimental** studies during **epidemics** is feasible?

If you DISAGREE or STRONGLY DISAGREE with one or more of the questions related to this outcome, please use this space below to provide your rationale

##### **Section 3: Overall acceptability of the COS**

In this section, we will ask you about the overall acceptability of the [COS](https://www.dropbox.com/scl/fi/5g91h52ui2bqgul528xq0/4.-Hyperlink_Box-1.docx?rlkey=k9yhbsftpx2rn5cq5t4rnkk8r&st=qnvknpo0&dl=0).

Please use the Likert scale options to answer.

* 36. To what extent do you agree that this COS captures the most important outcomes?

Neither Agree

Strongly Disagree	Disagree	Nor Disagree	Agree	Strongly Agree	*Don´t Know*

- 37. This COS is likely to be acceptable to key stakeholders.

Neither Agree

Strongly Disagree	Disagree	Nor Disagree	Agree	Strongly Agree	*Don´t Know*

- 38. To what extent do you agree that this COS will facilitate timely evidence generation for decision-making during outbreaks and epidemics?

Neither Agree

Strongly Disagree	Disagree	Nor Disagree	Agree	Strongly Agree	*Don´t Know*

- 39. How much effort will it take to use this COS?

Huge Effort	A Lot of Effort	Moderate Effort	A Little Effort	No Effort at All	*Don´t Know*

- 40. I intend to use this COS for maternal and neonatal health research and surveillance in the context of epidemics.

Strongly Disagree	Disagree

Neither Agree

Nor Disagree	Agree	Strongly Agree	*Don´t Know*

##### **Section 4: Barriers for COS adoption**

Adoption is a challenge for any well-developed COS. While several barriers have been reported in the literature, they primarily focus on clinical trials, with none specifically referring to observational studies.

41. Please, prioritize from the list below the potential barriers to the adoption of this particular COS, which focuses on maternal and neonatal health observational research and surveillance in the context of epidemics.

Poor knowledge and understanding of COS.

Lack of awareness of the COS.

COS not accessible in locally understandable languages. Difficulties/challenges choosing between multiple COS.

Lack of skills to apply the COS.

Lack of guidance and validated tools to collect data and measure outcomes.

Resource requirement associated with measuring the COS.

Concerns about quality of the COS development and methods for keeping it current.

Lack of perceived usefulness in generating timely evidence for decision-making.

COS could be seen as restrictive and limiting the range of outcomes.

Too many COS outcomes limits reporting of all outcomes.

Increased burden on researchers and patients due to the data collection required to report COS.

Researcher preference to choose own outcomes.

Lack of external incentives (e.g., recommendations by funders or regulatory agencies). Limited number of patient-centered outcomes.

None.

Other (please specify)

##### **Additional comments**

42. Please feel free to add any comments you consider pertinent to the overall acceptability of the COS or the barriers to its adoption.

##### **Thank you for your collaboration!**

Thank you for participating in this survey and for your continued support throughout the process for the development of this COS.

The involvement of a broad group of key stakeholders will significantly strengthen the COS, enhancing its applicability and validity across diverse settings. We greatly appreciate your time and interest.

As a token of our appreciation, we would like to offer you a certificate of participation. If you're interested, please email us at and we will arrange this for you.

43. We would also like to acknowledge your contribution in this survey report or publication. Please select the corresponding option:

Yes, I agree to be acknowledged in the report or publication of this survey No, I do not agree to be acknowledged
