## Supplementary File SF2: Additional Methods for "Acceptability and perceived barriers to adoption of the core outcome set for maternal and neonatal health research and surveillance during emerging and ongoing epidemic threats (MNH-EPI-COS). An online survey"

Summary of the methods of previous phases

**Final consensus on MNH-EPI-COS**

N

**MNH-EPI-COS DEVELOPMENT PROCESS**

**Systematic review**

N

**Consensus meetings** (online, in-person)

n = 24

N

**Delphi round 2** (online)

n = 141 / 150 (91 %)

N

**ACCEPTABILITY SURVEY**

**Online consultation**

n = 100/118 (85%)

- Acceptability of the final MNH-EPI-COS
- Acceptability of the definitions
- Anticipated barriers to COS adoption
- Perceived feasibility of data collection

**Delphi round 1** (online)

n = 150 / 197 invited (76%)

N
