## Supplementary File SF3: Participants of the online survey for "Acceptability and perceived barriers to adoption of the core outcome set for maternal and neonatal health research and surveillance during emerging and ongoing epidemic threats (MNH-EPI-COS). An online survey"

**Supplementary file SF3. Participants of the online survey who consented to be acknowledged**

Adejumoke Idowu Ayede

Albert Manasyan

Alberto Toso

Amy Boldosser-Boesch

Andy Stergachis

Ashraf Nabhan

Azeem Walele

Babagana Bako

Beate Kampmann

Brendan Carvalho

Carolina Carvalho Ribeiro Do Valle

Caroline Homer

Christina Ricci

Claire Thorne

Clare Whitehead

Cristina Cuesta

Daniela Noris Vasquez

David Kimberlin

Dorotheah Obiri

Eleni Vavouraki

Elhadi Miskeen

Eliana Marengo

Ellen O'keeffe

Erika Ota

Fleda M Jackson

Fook-Choe Cheah

France Donnay

Gabriela Tavchioska

Guilherme Amaral Calvet

Hellen Mutsi

Hemantha Senanayake

Hodorogea Stelian

Ilknur Okay

Jean Paul Ndayizeye

Jose Guilherme Cecatti

Jose Rojas

Joyce Browne

Julian Gustavo Antman

Katharina Hartmann

Ken Takahashi

Koiwah Koi-Larbi

Leah Greenspan

Liona Poon

Loic Sentilhes

Luigi Gagliardi

María Aurelia Giboin Mazzola

María J. Rodriguez-Sibaja

Maria Laura Costa Do Nascimento

Marleen Van Gelder

Meghan Azad

Michael G Gravett

Michal Lipschuetz

Michelle Sadler

Michelle Tan Hwee Pheng

Nasim Akhtar

Niveen Abu Rmeileh

Nzelle Delphine Kayem

Oleksandra Balyasna

Petr Velebil

Pierre Buekens

Pomar Léo

Priya Soma-Pillay

Raanan Raz

Redeat Workneh

Rhoda Amafumba

Ricardo Nieto

Rosemary Njogu

Rosnah Sutan

Safa Elhassan

Sarah Jorgensen

Satoru Ikenoue

Serge Alain Tougma

Shabina Ariff

Shivaprasad Goudar

Shuby Puthussery

Soledad Puppo

Sonia Deantoni

Soo Downe

Stephanie Dellicour

Tippawan Liabsuetrakul

Ursula Winterfeld

Wendy Pollock

Yousef Khader

Yuefang Huang

Zaleha Abdullah Mahdy
